# Assessing neuropathic pain in sickle cell disease: How useful is quantitative sensory testing?

**DOI:** 10.1101/2021.07.07.21260027

**Authors:** Zachary Ramsay, Damian Francis, Rachel Bartlett, Amza Ali, Justin Grant, Georgiana Gordon-Strachan, Monika Asnani

## Abstract

Quantitative sensory testing (QST) is a psychophysical test of sensory function which may assist in assessing neuropathic pain (NP). This study compares QST findings with a standardized NP questionnaire to assess their agreement among Jamaicans with sickle cell disease (SCD). A cross sectional study consecutively recruited SCD patients 14 years and older, not pregnant, and without history of clinical stroke or acute illness in Kingston, Jamaica. QST identified thresholds for cold detection, heat detection, heat pain and pressure pain at the dominant thenar eminence, opposite dorsolateral foot and the subject’s most frequent pain site. The Douleur Neuropathique 4 (DN4) was interviewer-administered to diagnose NP. Subjects were divided into low and high sensitization groups if below the 5^th^ and above the 95^th^ percentiles, respectively on QST measures. Kappa agreement coefficients, and receiver operator characteristic (ROC) curves were performed to compare QST with the DN4. Two-hundred and fifty-seven SCD subjects were recruited (mean age 31.7 ± 12.2 years, 55.7% female, 75% SS genotype). Kappa agreements were fair (0.2-0.4) to good (0.6-0.8) between DN4 individual items of itching, hypoesthesia to touch, hypoesthesia to pinprick and brush allodynia with various QST sensitization groups. However, kappa agreements between the NP overall diagnosis on the DN4 with sensitization groups were poor (<0.2). Only heat detection (0.75) and heat pain (0.75) at the leg as a pain site showed satisfactory area under the curve (>0.7). QST may assist in assessing individual components of NP but its use should be limited as a tool to augment clinical assessments.

## Introduction

Pain is the primary disease complication of sickle cell disease (SCD) and may be acute, chronic or neuropathic. The International Association for the Study of Pain defines neuropathic pain (NP) as “pain caused by a lesion or disease of the somatosensory system” (1), and is associated with positive symptoms, both evoked and spontaneous, as well as negative symptoms. Positive symptoms include gain of sensory function such as allodynia and hyperalgesia, and abnormal sensations or paraesthesia. Negative symptoms denote loss of sensation or hypoesthesia. NP has been found to occur in 25-40% of sickle cell disease (SCD) patients in previous studies (2), and is the result of peripheral and central sensitization, which can be measured by QST (3, 4). There is no true gold standard for the diagnosis of NP. Although NP can be assessed objectively using late laser evoked potentials and functional MRIs, these tests are expensive and not readily available.

Quantitative sensory testing (QST) and standardized questionnaires are both accessible and simple to conduct in large studies and clinical care. QST is a psychophysical test that assesses the function of sensory nerves and their pathways, based on the patient’s subjective response to standardized thermal, nociceptive or pressure stimulation. Thermal and pain stimuli assess small Aδ and C nerve fibres, while touch and vibration stimuli assess large Aα and Aβ fibres (5). Neuropathy, due to a disease or lesion of these sensory nerves, may result in significant loss or gain of sensory function that can be assessed by QST. The Douleur Neuropathique 4 (DN4) is a standardized questionnaire that has been validated to detect NP in patients with diabetic neuropathy (6). This has also been used in SCD with similar results to other NP tools (7). QST and DN4 have not been previously measured in the Jamaican SCD population.

QST has been measured in other SCD studies and compared to other NP questionnaires. The DN4 has been found to have the highest accuracy compared to other NP tools (8), however it has not been compared to QST in any SCD studies. For the first time, this study will assess the agreement of QST with the DN4 among Jamaicans with SCD. We also aimed to report the temperature and pressure values (known as ‘sensitization values’) at which patient perceive these QST stimuli, and the associated clinical and sociodemographic factors.

## Materials and Methods

A cross sectional study was done at the Sickle Cell Unit (SCU), which is an outpatient clinic in Kingston, Jamaica. The study was approved by the University of the West Indies Mona Campus Research Ethics Committee (ECP# 66, 17/18). Patients were consecutively recruited if they were 14 years of age or older, with no active acute illness, no history of clinical stroke and not pregnant. Written informed consent (including child assent and parental/guardian consent for those under 16 years of age) was obtained from all patients prior to study procedures. In order to estimate at least 25% of SCD patients with NP (9), based on 2120 patients over the age of 14 years old with at least 1 well SCU clinic visit in 2017, 5% margin of error, and 5% significance level, we would need to recruit at least 254 subjects.

### Questionnaire instruments

Subjects completed a study instrument with sociodemographic, clinical information and medication history. The Pain Episode module of the Adult Sickle Cell Quality of Life Measurement Information System questionnaire was completed to assess vaso-occlusive pain as a composite ‘acute pain score’. This module has a total possible score of 33 from 5 items, where higher scores indicate higher frequency and severity. Subjects were classified as using strong opioids if they reported morphine or pethidine use in the last 4 weeks. Anthropometric measures were also recorded, and genotype was categorized as severe when subjects were SS or Sβ0 thalassemia and mild when SC or Sβ+ thalassemia.

### Douleur Neuropathique 4 (DN4) questionnaire

DN4 questionnaire was interviewer administered to diagnose “likely NP”. The DN4 contains seven NP descriptors, and three examination items that assess for brush allodynia and hypoesthesia to touch and pinprick. Each positive finding has a binary score, and NP is assessed with a total score of at least 4 out of 10.

### Quantitative sensory testing

The method of limits was performed as described previously by Rolke et al (10), on the dominant thenar eminence and opposite dorsolateral foot. Subjects also indicated the body part where they feel pain most often, and this was tested whenever it was identified. When patients had a leg ulcer, the side, whether left or right, of the thenar eminence and lateral foot tested were switched, so that the foot with the ulcer was not tested. Cold detection threshold (CDT), heat detection threshold (HDT) and heat pain threshold (HPT) were measured using the Q-Sense instrument (Medoc Ltd). Pressure pain threshold (PPT) was also measured using an algometer, the AlgoMed (Medoc Ltd). Each stimulus was applied starting from a specific baseline and then decreased or increased at a specific rate until the subject indicated that they detected the stimulus with CDT and HDT tests, and then separately when it became painful with HPT and PPT tests. For cold and heat stimulation, a baseline of 32°C, rate of 1°C/second, minimum of 16°C and maximum of 50°C were used. We used a rate of 35kPa/second for PPT. All measurements were repeated three times and the average value was recorded.

### Statistical methods

CDT and HDT were calculated by subtracting the temperature indicated by the subject from the baseline, whereas HPT and PPT were recorded as the actual value indicated. These were described using medians and interquartile ranges, and then compared to clinical and sociodemographic covariates using Wilcoxon rank sum, Kruskal-Wallis and Spearman’s rank order correlation tests. The association of these continuous QST values were compared with the binary variables of the overall outcome of the DN4 using Wilcoxon rank sum tests. Bootstrapped receiver operating characteristic (ROC) curves with 1000 repetitions were also performed to assess the area under the curve (AUC) when the binary NP outcome of the DN4, as the reference predictor, was compared with the continuous QST values. We interpreted ROC area under the curve (AUC) values as follows: unacceptable (<0.7), acceptable (0.7-0.8), excellent (0.8-0.9), and outstanding (>0.9) (11).

QST values for each modality were also categorized into binary outcomes of low sensitization if below the 5^th^ percentile for the sample and high sensitization if above the 95^th^ percentile, respectively. Agreements between these binary variables and both the overall DN4 binary outcome and the individual items were assessed using kappa agreement coefficient. We interpreted the kappa statistic as follows: poor (<0.2), fair (0.2-0.4), moderate (0.4-0.6), good (0.6-0.8), and very good (0.8-1.0) (12). The internal consistency of the DN4 was evaluated using Cronbach’s alpha coefficient.

Given the ethnic limitations of available QST reference values at the time of the study, we calculated z-scores for subjects 18 years or older based on the protocol of the German Research Network on Neuropathic Pain (10) as a secondary sensitivity analysis. Z-scores were calculated for HDT, HPT and CDT at the thenar eminence and dorsolateral foot, and PPT at the thenar eminence only. These continuous z-score values were compared to binary DN4 outcomes of the overall questionnaire and each individual item using Wilcoxon rank sum tests. ROC curve analysis was also used to compare z-scores with overall DN4 binary outcomes. All analyses were performed in Stata 14.2.

## Results

There were 257 subjects recruited with mean age of 31.7 ±12.2 years, 55.7% female and 75% homozygous SS genotype, including 80.5% severe, 19.5% mild.

### Clinical factors

There were 221 persons who reported having a pain site where they experienced pain most frequently, of which the lower back (22.3%) and leg (14.5%) were most common. There were 23 persons who reported hydroxyurea use (9.0%), 24 who used strong opioids (13.2%) and 35 with current leg ulcers (13.6%). Body mass index (BMI) was distributed as follows: underweight (59, 23.1%), normal (142, 55.7%), overweight (39, 15.3%), obese (15, 5.9%). There was a mean acute pain score of 18.7±7.5 ranging from 0-31. The internal consistency coefficient for the DN4 was 0.735, and 25.7% were assessed as having NP based on a score of 4 out of 10.

### Quantitative sensory testing

Thresholds for low and high sensitization for each QST modality is shown in Table 1. Subjects were most sensitive to CDT and HDT at the thenar eminence, HPT at the lower back and PPT at the legs (Table 2).

**Table 1.** Low and high sensitization thresholds among Jamaicans with sickle cell disease

|  | Thenar eminence |  |  |  | Dorsolateral foot |  |  |  | Back |  |  |  | Legs |  |  |  |
| --- | --- | --- | --- | --- | --- | --- | --- | --- | --- | --- | --- | --- | --- | --- | --- | --- |
|  | CDT | HDT | HPT | PPT | CDT | HDT | HPT | PPT | CDT | HDT | HPT | PPT | CDT | HDT | HPT | PPT |
| L | -5.3 | 0.8 | 35.2 | 164.3 | -9.1 | 1.5 | 35.8 | 144.1 | -6.7 | 1.1 | 35.3 | 122.2 | -8.5 | 1.5 | 36.6 | 41.5 |
| H | -0.7 | 4.6 | 47.6 | 664.4 | -0.9 | 12.2 | 47.7 | 544.2 | -1.5 | 7.2 | 46.2 | 691.2 | -0.7 | 13.3 | 47.4 | 699.1 |

L: low sensitization threshold which is 5<sup>th</sup> percentile; H: high sensitization threshold which is the 95<sup>th</sup>
percentile; CDT: cold detection threshold (°C), HDT: heat detection threshold (°C), HPT: heat pain threshold (°C), PPT: pressure pain threshold (kPa)
Sensitization values are calculated by subtracting the temperature identified by the patient from a baseline of 32°C

**Table 2.** Quantitative sensory testing values among Jamaicans with sickle cell disease

|  | Thenar eminence | Lateral foot | Lower back | Legs |
| --- | --- | --- | --- | --- |
| CDT (°C) | -1.8 (-2.8, -1.2) | -2.5 (-4.2, -1.5) | -2.4 (-4, -1.5) | -2.4 (-4, -1.5) |
| HDT (°C) | 1.6 (1.2, 2.3) | 4.0 (2.6, 6.8) | 3.2 (2.3, 4.6) | 6.5 (4.3, 9.5) |
| HPT (°C) | 41.1 (38.1, 43.7) | 41.0 (38.3, 44.2) | 39.0 (37.0, 42.1) | 42.6 (39.6, 45.5) |
| PPT (kPa) | 320.3 (240.9, 419.4) | 283.9 (202.2, 380.6) | 317.9 (221.8, 428.8) | 267.9 (167.3, 378.0) |

Values shown are the median (interquartile range); CDT: cold detection threshold, HDT: heat
detection threshold, HPT: heat pain threshold, PPT: pressure pain threshold
Cold and heat detection thresholds are calculated by subtracting the temperature identified by the patient from a baseline of 32°C

### Clinical and sociodemographic associations of QST values

Older persons were less sensitive to HDT (rs=0.2, P=0.004) at the thenar eminence, and CDT, HDT and HPT at the dorsolateral foot (Table 3). Similarly, persons with higher acute pain scores were more sensitive to PPT at both the thenar eminence. Females were more sensitive to all modalities except CDT at the thenar eminence. Subjects with leg ulcers were more sensitive to all modalities at the dorsolateral foot except PPT, while those currently using hydroxyurea were more sensitive to HDT at the thenar eminence. Subjects who used strong opioids in the last 4 weeks were more sensitive to PPT at the thenar eminence.

**Table 3.** Significant clinical and sociodemographic associations of quantitative sensory testing sensitization thresholds among Jamaicans with sickle cell disease

| Thenar eminence sensory modalities |  |  |  |  |
| --- | --- | --- | --- | --- |
| Covariates: | CDT | HDT | HPT | PPT |
| Age, correlation (*P*)<sup>1</sup> | -0.111 (0.078) | 0.178 (0.004) | 0.040 (0.039) | 0.021 (0.741) |
| Sex<sup>2</sup>, *P* | 0.96 | 0.009 | <0.0001 | <0.0001 |
| Male, median (IQR) | -1.8 (-2.8, -1.3) | 1.74 (1.3, 2.6) | 42.2 (39.1, 45.5) | 377.6 (300.2, 497.2) |
| Female, median (IQR) | -1.7 (-3.0, -1.2) | 1.4 (1.1, 2.1) | 39.8 (37.7, 42.6) | 277.0 (215.6, 367.2) |
| Hydroxyurea use<sup>2</sup>, *P* | 0.387 | 0.034 | 0.691 | 0.212 |
| Present, median (IQR) | -1.6 (-2.1, -1.0) | 1.2 (1.1, 1.6) | 41.1 (38.0, 44.7) | 358.0 (274.1, 459.3) |
| Absent, median (IQR) | -1.8 (-2.9, -1.3) | 1.6 (1.2, 2.4) | 41.0 (38.1, 43.7) | 316.9 (239.1, 411.6) |
| Strong opioids<sup>2</sup>, *P* | 0.90 | 0.51 | 0.95 | 0.02 |
| Present, median (IQR) | -1.7 (-2.9, -1.3) | 1.6 (1.3, 2.3) | 41.9 (38.0, 43.0) | 287.0 (189.1, 349.2) |
| Absent, median (IQR) | -1.8 (-2.8, -1.7) | 1.6 (1.1, 2.3) | 40.7 (38.1, 43.9) | 328.0 (243.4, 431.5) |
| Acute pain score, correlation (*P*)<sup>1</sup> | 0.009 (0.883) | 0.056 (0.371) | -0.067 (0.287) | -0.134 (0.033) |
| Dorsolateral foot sensory modalities |  |  |  |  |
| Covariates: | CDT | HDT | HPT | PPT |
| Age, correlation (*P*)<sup>1</sup> | -0.24 (<0.001) | 0.25 (<0.001) | 0.278 (<0.001) | -0.060 (0.336) |
| Sex<sup>2</sup>, *P* | 0.004 | <0.0001 | <0.0001 | <0.0001 |
| Male, median (IQR) | -3.0 (-4.7, -1.8) | 5.2 (3.1, 8.9) | 42.6 (38.8, 44.8) | 350.8 (256.4, 424.3) |
| Female, median (IQR) | -2.4 (-3.5, -1.4) | 3.4 (2.3, 5.5) | 40.6 (38.2, 44.0) | 252.8 (186.9, 314.3) |
| Presence of leg ulcers<sup>2</sup>, *P* | 0.012 | 0.0001 | 0.012 | 0.336 |
| Present, median (IQR) | -2.9 (-8.4, -2.1) | 6.2 (3.6, 11.5) | 41.7 (39.9, 46.8) | 312.9 (199.9, 422.4) |
| Absent, median (IQR) | -2.5 (-4.0, -1.5) | 3.8 (2.5, 6.0) | 40.7 (38.1, 43.9) | 279.3 (204.5, 377.6) |

Statistical tests: 1: Spearman's correlation 2: Wilcoxon Rank-sum test. Covariates that had no significant associations or correlations were not included in the table.

### Comparing QST with DN4

Subjects assessed as having likely NP on the DN4 were less sensitive to HDT at the thenar eminence (P=0.02), CDT (P=0.007) and HDT (P=0.02) at the dorsolateral foot, and HDT (P=0.01) and HPT (P=0.01) at the leg. Kappa agreements were fair (0.2-0.4), moderate (0.4-0.6) and good (0.6-0.8) between various items of the DN4 and QST sensitization groups (Table 4). Other kappa agreements between low and high sensitization groups were poor (<0.2) when compared with the individual items and the overall outcome.

**Table 4.** Fair to good kappa agreements between individual items of the DN4 and groups of hyposensitive and hypersensitive Jamaicans with sickle cell disease

|  | Dorsolateral Foot |  | Back |  | Leg |
| --- | --- | --- | --- | --- | --- |
|  | CDT<sub>low</sub> | HDT<sub>high</sub> | HDT<sub>high</sub> | HPT<sub>low</sub> | PPT<sub>high</sub> |
| Itching | 0.21*** | 0.07 | -0.05 | 0.79 | 0.09 |
| Hypoesthesia touch | 0.27*** | 0.27*** | 0.37** | -0.06 | 0.16 |
| Hypoesthesia pinprick | 0.33*** | 0.21*** | 0.21* | 0.79*** | 0.12 |
| Brush allodynia | -0.02 | -0.02 | 0.00 | 0.00 | 0.48*** |

P<0.05: \*, P<0.01: \*\*, P<0.001:\*\*\*; CDT<sub>low</sub>: low sensitization cold detection group; HDT<sub>high</sub>: high
sensitization heat detection group; HDT<sub>low</sub>: low sensitization heat detection group; HPT<sub>high</sub>: high
sensitization heat pain group; PPT<sub>high</sub>: high sensitization pressure pain group; Domains that only had
poor kappa agreements (<0.2) have been excluded from this table

**Table 5.**
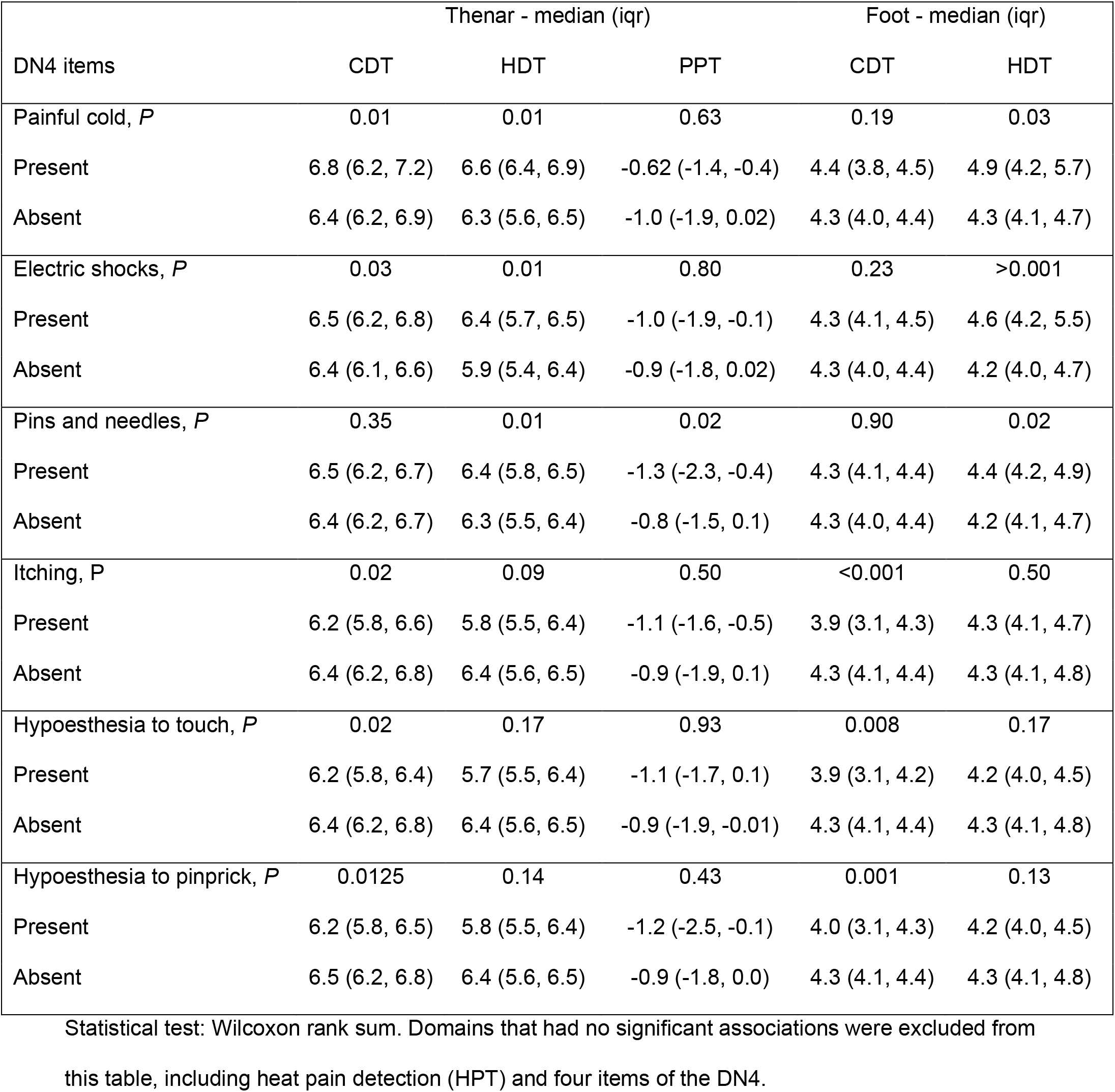
Associations of DN4 items with z-scores QST sensitization values based on normative values among Jamaicans with sickle cell disease

### Receiver operator characteristic (ROC) curves

Acceptable AUC (0.7-0.8) were only with ROC curves assessing HDT (0.75) and HPT (0.75) at the legs using the DN4 as the reference standard (Figure 1).

**Figure 1.**
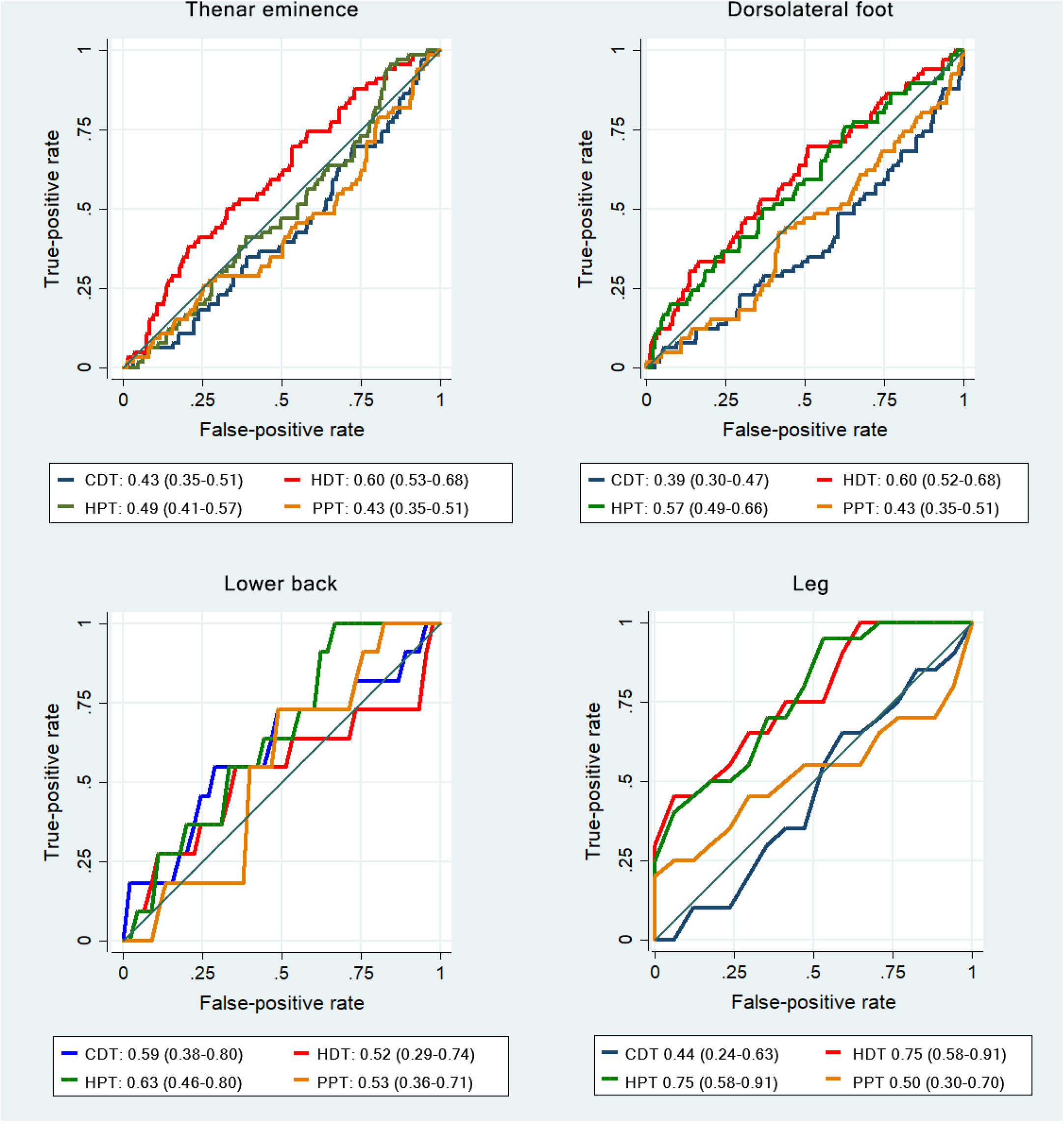
Receiver Operating Characteristic Curves assessing thermal and pressure quantitative sensory testing values as predictors of neuropathic pain using the Douleur Neuropathique 4 as the reference standard. CDT: cold detection, HDT: heat detection, HPT: heat pain threshold, PPT: pressure pain threshold. Wald tests if classifiers have equal AUC values: thenar eminence (P=0.005), dorsolateral foot (P=0.001), back (P=0.617), leg (P=0.086).

### Sensitivity analysis

The associations of the z-scores of thermal sensitization values of subjects 18 years of age or older compared with the DN4 are shown in Table 4. Comparatively, all ROC curves were unacceptable (<0.7).

## Discussion

We aimed to evaluate the agreement of QST and the DN4 in assessing NP in Jamaicans with SCD and found fair to good agreements and associations with some items of the questionnaire. However, agreements with the overall NP rating using the DN4 questionnaire were poor. Although QST is useful in assessing the negative and positive evoked sensory symptoms of NP, it does not assess other components such as spontaneous pain. For these reasons, QST is best used as a diagnostic tool for small fibre neuropathy, and should only be used to augment clinical assessments of NP (5).

Agreements and AUC were higher at leg and back pain sites, compared to the thenar eminence and foot. This may suggest that although abnormal sensitization is often found at the peripheries in SCD (13), which is where most investigators perform QST when assessing for NP, it is likely best to perform QST at the specific painful sites. This is a limitation of QST as reference ranges are not available for all possible sites, and it would be difficult to assess many different pain sites collectively in one study. Notably, QST measurements also did not agree well with questionnaire items that seem similar in description. For example, sensations of burning had poor agreement with HDT and HPT. This highlights the disconnect between pain perception and actual nociceptive stimuli in NP, which results from aberrant and spontaneous pain signals. Also, questionnaire items that involve examination do not agree with QST measurements that may seem similar because they are assessing the function of different nerve fibres. For example, an examination for hypoesthesia to light touch relates to Aβ fibres whereas QST pressure pain, from prolonged mechanical touch, relates to C fibres. Similar to these findings, QST has been unable to differentiate persons with painful diabetic neuropathy from those with painless neuropathy (14, 15). However, Dyal et al (16) found that SCD subjects classified as sensitized on QST (based on the author’s diagnostic decision tree) scored higher on the Neuropathic Pain Symptom Inventory and a weighted score of the PAINReportIt questionnaire.

We also found that females were more sensitive to all modalities at the thenar eminence and dorsolateral foot, and that older subjects less sensitive to all at those sites except HPT and PPT at the thenar eminence and PPT at the dorsolateral foot. Sex differences are not commonly seen in QST in SCD studies (17, 18). However, Brandow et al (13) have reported that both older SCD subjects and older non-SCD controls were more sensitive to heat and mechanical stimuli and that although there were no overall significant sex differences between groups, females with SCD had higher mechanical pain thresholds compared to males. In this study, those who used hydroxyurea and strong opioids were more sensitive to HDT and PPT at the thenar eminence, respectively. This is the opposite to previous findings where SCD subjects on hydroxyurea had higher heat and mechanical pain thresholds compared to SCD subjects not on hydroxyurea (19), but similar to findings that chronic opioid therapy is associated with central sensitization in SCD (20). Higher acute pain scores were associated with being more sensitive to PPT at both the thenar eminence and dorsolateral foot. This is consistent with the underlying pathogenesis of NP in SCD. Recurrent vaso-occlusive crises result in ongoing and persistent nociceptive insults, leading to peripheral hyperexcitability and sensitization, and then also neurogenic inflammation in the central nervous system with resultant additional central sensitization (3, 4). In this regard, we indeed found that the subset of subjects with leg ulcers were more sensitive to thermal stimuli at the opposite dorsolateral foot, indicating a diffuse pattern of thermal sensitization. This is a novel QST finding in SCD and could suggest that the development of peripheral neuropathy may be one of the early steps leading to an increased propensity of some SCD patients to developing leg ulcers. This may itself involve several downstream mechanisms including neurogenic inflammation and trophic alterations to the skin from autonomic small fibre alterations affecting the skin microvasculature and the eventual development of leg ulcers in SCD (21).

In summary, quantitative sensory testing does not replace the requirement for a careful history, clinical examination and validated tools such as the DN4 in identifying a potential neuropathic pain syndrome in this patient population who otherwise have many causes of pain. QST may however be used to assess small fibre neuropathy and in specific instances this may help to increase diagnostic rigour. Peripheral nerve conduction studies and intraepidermal nerve fibre density studies are needed to further assess, respectively, the contribution of large fibre neuropathy and small nerve fibre structure, to better understand this unique sensory phenotype. The implications of accurate diagnosis are significant as the treatment modalities for treating neuropathic pain are quite different from those generally used in SCD pain management. Furthermore, the potential impact of ineffective treatment on the patient’s long-term pain trajectory is profound. While certain tentative characteristics have been identified in this study, such as gender and age, more work is required to confirm the demographic and clinical characteristics of patients most at risk of neuropathic pain.

### Limitations

We excluded persons with history of clinical stroke because we expect them to have sensory changes that would confound with the changes that we expect in SCD from central sensitization. The exclusions, which also included persons with acute active illness, may have resulted in us recruiting subjects with a milder phenotype.

This was created using Stata 14.2.

## Supporting information

STROBE checklist

## Data Availability

Data may be available upon request

## Acknowledgments

We would like to thank Ofrit Bar-Bachar of Medoc Ltd for her review of the methodology.

## Declarations

### Availability of data and material

Data may be available upon request

### Code availability

Not applicable

### Author contributions

ZR was involved with conceptualization, data collection, data analysis and writing the manuscript

DF was involved with conceptualization and writing the manuscript

RB was involved with conceptualization, data collection and writing the manuscript

AA was involved with conceptualization and writing the manuscript

JG was involved with conceptualization and writing the manuscript

GGS was involved with conceptualization and writing the manuscript

MA was involved with conceptualization, data analysis and writing the manuscript

